# Development and internal validation of risk scores to predict survival in the pediatric population following out-of-hospital cardiac arrest

**DOI:** 10.1101/2025.08.04.25332995

**Authors:** Minaz Mawani, Bryan McNally, Jessica Knight, Ye Shen, Mark Ebell

**Affiliations:** Larner College of Medicine, University of Vermont Department of Medicine, Burlington, Vermont; Epidemiology and Biostatistics, University of Georgia College of Public Health, Athens, Georgia; Emory University School of Medicine, Atlanta, Georgia; Department of Family Medicine, Michigan State University, East Lansing, Michigan

**Keywords:** Out-of-hospital cardiac arrest, Pediatric population, risk scores, clinical decision-making

## Abstract

**Introduction:** Out-of-hospital cardiac arrest (OHCA) in the pediatric population is associated with poor survival and neurological outcomes. We aimed to develop and internally validate risk scores to predict survival to discharge in pediatric OHCA patients.

**Methods:** We included pediatric OHCA patients in the Cardiac Arrest Registry to Enhance Survival from 2013 to 2023. We used logistic regression (LR) and classification and regression trees (CART) to develop risk scores using 70% of the data and validated them using the remaining 30% of the data. We used discrimination and predictive accuracy to evaluate model performance.

**Results:** We included 26895 pediatric patients, of whom 27.8% survived to hospital admission and 11.9% survived to hospital discharge. We developed three separate risk scores for i) infants ii) preschool and school-age children, and iii) adolescents. The most important predictor variables were gender, arrest location, witness status, etiology of arrest, bystander CPR, use of prehospital AED, and first rhythm. Models developed using both LR and CART approaches had good classification accuracy ( AUROCC for LR: 0.81, 0.77 and 0.82, AUROCC for CART: 0.78, 0.76, 0.82 for the three age groups) However, we found CART models to be the most useful because they are simpler to follow, classified more patients in low and high survival categories using a smaller number of predictors. The average survival probability in infants for each risk group was 3%, 13%, and 33%, for preschool and school-age children it was 4%, 11%, and 29%, whereas for adolescents it was 4%, 13%, and 48% for the low, moderate, and high survival categories.

**Conclusion:** Pediatric patients experiencing OHCA can be classified into low, moderate and high survival categories using a simple risk score and easily identified prehospital variables. These risk scores can facilitate research, monitor the quality of medical services and can potentially support clinical decision-making.

## Introduction

Out-of-hospital cardiac arrest (OHCA) in the pediatric population is an uncommon event, associated with poor survival and neurological outcomes ^1^. Despite ongoing efforts to improve outcomes, less than 10% survive to hospital discharge and many children have severe neurological deficits ^2 3 3 4^. Several factors predict the outcomes of OHCA in children including age, specific comorbid conditions, no-flow time, etc. ^1,5–7^. In the absence of any prediction models, physicians’ prediction of survival and functional outcomes have been observed to be inaccurate ^8,9 10^. Predicting survival outcomes can potentially help guide decision-making for individual patients ^11^.

Risk scores, also known as prediction models or clinical prediction rules, combine multiple predictors such as patient characteristics, laboratory results, comorbid conditions, and other variables to estimate the probability of a clinical outcome ^12 13–15^. When appropriately developed and validated, risk scores have several potential advantages over clinical judgment alone. These models can provide objective and reliable results with high accuracy while taking several factors into account as compared to clinical judgment alone which can sometimes be inconsistent and biased^16^. Several risk scores to predict survival outcomes of OHCA in adults have been developed and validated ^17–19^. However, these models cannot be applied to the pediatric population because of differences in etiology, comorbidities, presenting rhythms, and outcomes^20,21^.

A model to predict OHCA outcomes in pediatric patients has been developed and validated in Japanese population using prehospital variables ^22^. The study used older data, estimated predictors of one-month survival, used limited pre-hospital variables and was not externally validated to be used in a different population.

Therefore, we aimed to develop and internally validate a risk score to predict survival to discharge in pediatric out-of-hospital cardiac arrest patients with ROSC (return of spontaneous circulation) who survived to ED admission using data on pre-hospital variables from 2013 to 2023. This score has the potential to inform physicians regarding prognosis and provide a tool for evidence-based decision-making to complement their clinical judgment.

## Methods

### Study design

In this retrospective study, deidentified data for model development and validation were obtained from patients in the CARES prospective registry from 2013 to 2023. The study was approved by the institutional review board (IRB) at the University of Georgia, and written informed consent was waived owing to the use of de-identified data from the registry. CARES registry data management system is secure and confidential, and the data is available only to authorized users. To maintain confidentiality, all data is shared in a de-identified, aggregate format. Data sharing applications and agreements are proposal-specific and limited to each site ^23^.

### Data sources

CARES is currently the largest out-of-hospital cardiac arrest registry in the United States. Data collection is based on the Utstein-style definitions-a standardized template of uniform reporting guidelines for clinical variables and patient outcomes that was developed by international resuscitation experts. The CARES web-based software (https://mycares.net), links three sources to describe each OHCA event: 1) 911 call center data, 2) EMS data, and 3) hospital data ^23,24^. Since its inception, the registry has been expanded and it now includes about 40 states, representing a catchment area of approximately 150 million people, about 46% of the US population. Throughout the country, more than 2000 EMS agencies and 2500 hospitals participate in this registry ^23^. Several measures are taken to ensure the integrity and accuracy of the data. These measures include standardized training of all CARES users, built-in software logic, an audit algorithm ensuring consistent data validation across the registry, and a bi-annual assessment of population coverage and case ascertainment.

### Population

Pediatric patients (≤ 18 years) experiencing a non-traumatic out-of-hospital cardiac arrest, captured in the CARES registry from 2013 to 2021 and surviving to the ED admission were included. Arrests that did not involve 911, were not treated, achieved a return of spontaneous circulation without defibrillation or CPR by 911 responder and those with missing data on outcomes were excluded.

### Predictor and outcome variable definitions

Standardized operational definition of the variables were used from the most recent CARES data dictionary. Potential variables included a) demographics such as age, gender, race/ethnicity, b) arrest-related characteristics such as date of arrest, location of arrest, witness status, etiology of arrest, first monitored rhythm, ROSC, and sustained ROSC, c) prehospital care information such as resuscitation attempted, bystander CPR provision, dispatcher CPR instructions, application of AED (automated external defibrillator), defibrillation, use of hypothermia, return of spontaneous circulation and pre-hospital survival status d) hospital-related variables such as emergency department outcome, survival to hospital admission, survival to hospital discharge, neurological outcome on discharge, diagnosis on discharge, and e) response and treatment times such as time of dispatch, time of arrival on the scene, and time from arrest to first CPR. The primary outcome was survival to hospital discharge.

### Model development

The conduct and reporting of this study is in accordance with the transparent reporting of a multivariable prediction model for individual prognosis or diagnosis (TRIPOD) guideline^25^. After checking for duplicates, item analysis and data visualization were used to assess the reliability and validity of the selected variables. New variables were defined as required by combining categories with small numbers. Missing values and outliers in the data were further explored with the CARES data team. Cases with missing information on the outcome were excluded from the analysis.

Categorical variables are presented as numbers and percentages, and continuous variables are presented as means ± standard deviation (SD). The significance of differences between groups with or without a risk factor for survival was determined by the chi-square test or independent sample T-test. A two-sided P value of less than 0.05 was considered to be statistically significant. For multivariable analysis we excluded variables exhibiting multicollinearity or with a very low prevalence. We used complete case analysis for variables with <5% missing data.

We developed separate models for infants (< 1 yr), preschool and school age children (1 to 12 years) and adolescents (13 to 18 years) following CARES guidelines. The data was divided into development (70%) and validation (30%) sets using simple random sampling. We compared two different modeling approaches: logistic regression (LR), and classification and regression trees (CART).

#### Logistic regression to derive point score-based risk prediction models

All analyses were carried out using SAS 9.4 (SAS Institute Inc, Cary, North Carolina, USA). We used LASSO (least absolute shrinkage and selection operator) to identify a simpler model with fewer, most suitable predictors of survival to build the model. A P-value <0.05 was considered statistically significant. We report adjusted odds ratios with 95% confidence intervals. We developed point scores by dividing the beta-coefficients by the smallest beta coefficient and rounding the result. The rounded beta-coefficients from the final model were used to construct the score.The scores were divided into low, moderate, and high likelihood of survival categories^26,27^

#### Classification and regression trees (CART) based prediction models

All CART analyses were carried out using JMP Pro 17.2.0 (SAS Institute Inc, Cary, North Carolina, USA). Classification and regression tree (CART) analysis was used to develop a model predicting survival outcomes. The algorithm uses non-parametric tests and recursive partitioning to evaluate data and progressively classify patients into subgroups based on the optimal independent predictors. The best predictor variable is determined by calculating the likelihood ratio chi-square test for each possible split. The variables and discriminatory values used and the order in which the splitting occurs are produced by the underlying mathematical algorithm and are calculated to maximize the resulting predictive accuracy. The algorithm identifies the predictor variable that best discriminates between patients with and without the outcome. We considered all important variables for the CART models. To optimize the performance of the model for future data and avoid overfitting, a combination of stopping rules and pruning were applied to the tree. We restricted that a minimum of 50 observations be allowed in any terminal node. The unsupervised modeling approach automatically selected the splitting variable based on the log worth statistic and continued until there was no improvement in the AUROCC (area under the receiver operating characteristic curve). Ends with similar probabilities of the outcome were grouped to form patient groups with low, moderate, and high probability of survival to discharge.

### Assessment of model performance

Model performance for the two methods (LR and CART) was evaluated using discrimination (area under the receiver operating characteristic curve or AUROCC) and the ability of the model to classify patients in low, moderate and high survival categories. One goal was to have as many patients as possible in the low and high risk groups to best aid clinical decisions. Results were compared to find the simplest model with optimum performance.

## Results

### Study population

Our analytic sample included data on 26895 children (≤ 18 years) experiencing OHCA. Almost half of the included participants were infants (48%), and 59% were males. Characteristics and outcomes of the study population are described in Table 1. Most of the arrests occurred in place of residence (85.7%), and 69.5% were unwitnessed. Almost 45% of the patients received bystander CPR, and only 7% of all patients had a shockable rhythm. Only 11.9% of children survived to hospital discharge.

**Table. 1:**
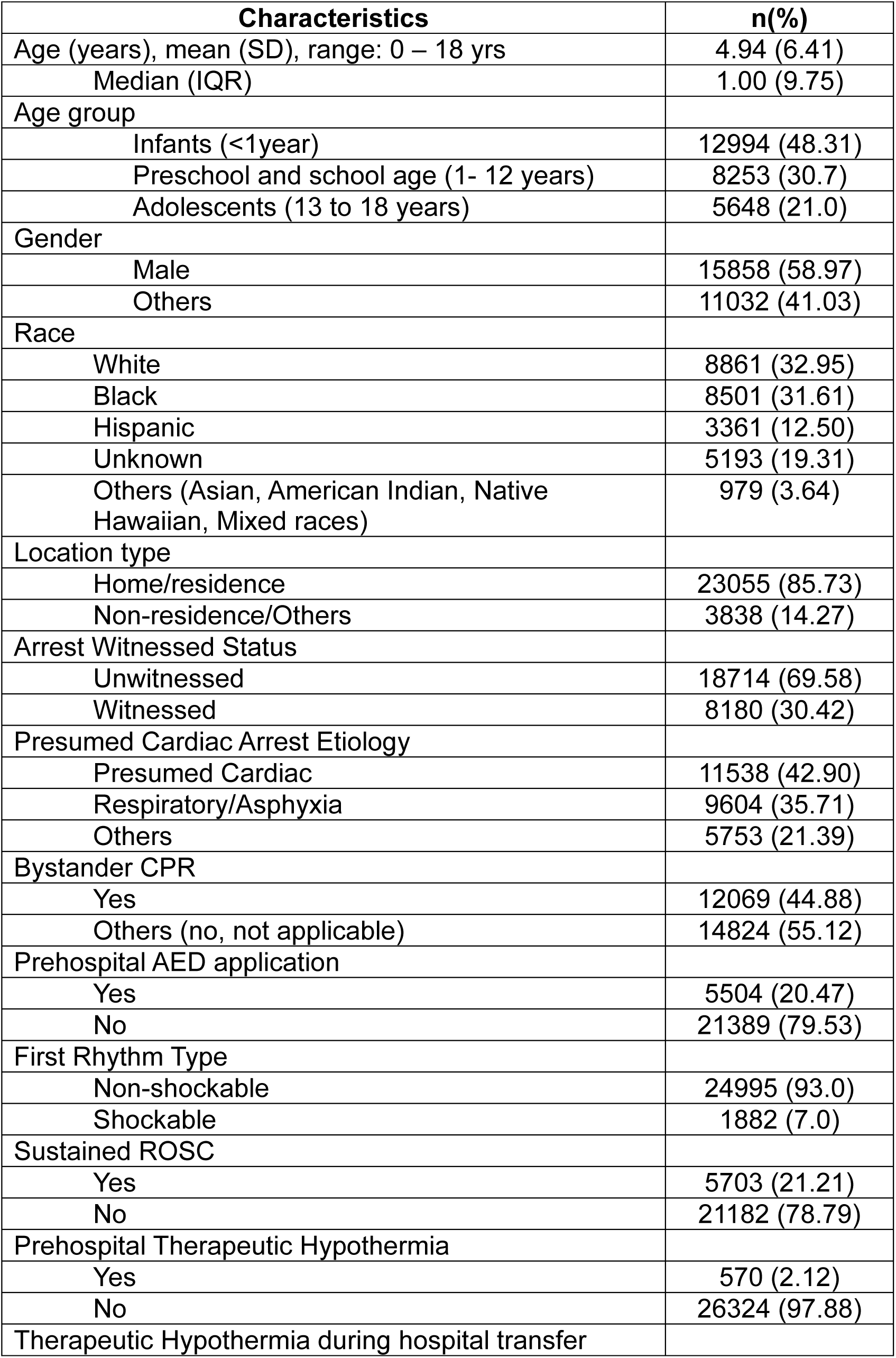
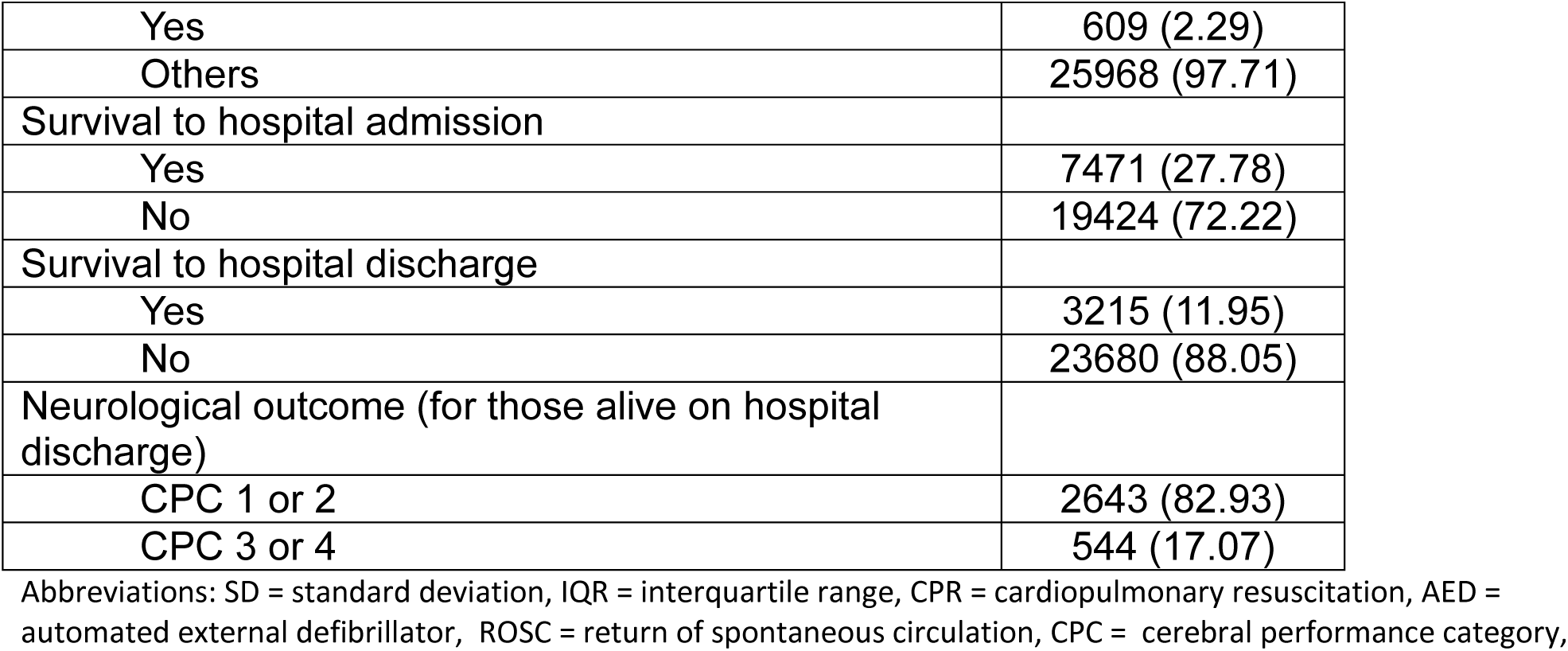
Characteristics of the study participants (n = 26895)

| Characteristics | n(%) |
| --- | --- |
| Age (years), mean (SD), range: 0 – 18 yrs | 4.94 (6.41) |
| Median (IQR) | 1.00 (9.75) |
| Age group |  |
| Infants (<1year) | 12994 (48.31) |
| Preschool and school age (1- 12 years) | 8253 (30.7) |
| Adolescents (13 to 18 years) | 5648 (21.0) |
| Gender |  |
| Male | 15858 (58.97) |
| Others | 11032 (41.03) |
| Race |  |
| White | 8861 (32.95) |
| Black | 8501 (31.61) |
| Hispanic | 3361 (12.50) |
| Unknown | 5193 (19.31) |
| Others (Asian, American Indian, Native Hawaiian, Mixed races) | 979 (3.64) |
| Location type |  |
| Home/residence | 23055 (85.73) |
| Non-residence/Others | 3838 (14.27) |
| Arrest Witnessed Status |  |
| Unwitnessed | 18714 (69.58) |
| Witnessed | 8180 (30.42) |
| Presumed Cardiac Arrest Etiology |  |
| Presumed Cardiac | 11538 (42.90) |
| Respiratory/Asphyxia | 9604 (35.71) |
| Others | 5753 (21.39) |
| Bystander CPR |  |
| Yes | 12069 (44.88) |
| Others (no, not applicable) | 14824 (55.12) |
| Prehospital AED application |  |
| Yes | 5504 (20.47) |
| No | 21389 (79.53) |
| First Rhythm Type |  |
| Non-shockable | 24995 (93.0) |
| Shockable | 1882 (7.0) |
| Sustained ROSC |  |
| Yes | 5703 (21.21) |
| No | 21182 (78.79) |
| Prehospital Therapeutic Hypothermia |  |
| Yes | 570 (2.12) |
| No | 26324 (97.88) |
| Therapeutic Hypothermia during hospital transfer |  |
| Yes | 609 (2.29) |
| Others | 25968 (97.71) |
| Survival to hospital admission |  |
| Yes | 7471 (27.78) |
| No | 19424 (72.22) |
| Survival to hospital discharge |  |
| Yes | 3215 (11.95) |
| No | 23680 (88.05) |
| Neurological outcome (for those alive on hospital discharge) |  |
| CPC 1 or 2 | 2643 (82.93) |
| CPC 3 or 4 | 544 (17.07) |

Abbreviations: SD = standard deviation, IQR = interquartile range, CPR = cardiopulmonary resuscitation, AED = automated external defibrillator, ROSC = return of spontaneous circulation, CPC = cerebral performance category,

### Bivariate analysis

We developed separate models for infants (<1 year), preschool and school age children (1 – 12 years), and adolescents (13 to 18 years) because of differences in characteristics, comorbid conditions, initial rhythms, and survival outcomes across these three populations. In bivariate analysis, male gender, witnessed arrest, public location of arrest, and shockable rhythm were associated with better survival in all age groups. Having a respiratory etiology of arrest was associated with better survival in infants. In contrast, in preschool and school-age children, etiology other than cardiac or respiratory was associated with better survival, and in adolescents, cardiac etiology was associated with better survival. AED application was associated with better survival in adolescents only. Receiving bystander CPR was associated with better survival in preschool, school-age and adolescents. Having an African American race was associated with poor survival in all age groups. Tables S1, S2, and S3 show characteristics of participants and factors associated with survival in the three age groups. Prehospital therapeutic hypothermia was used in <3% of the study population and was not considered for further analysis. Race was not included in the model since differences in the outcomes represent racial disparities rather than biological differences.

### Logistic regression models

Tables S4, S5 and S6 describe the results of the multivariable logistic regression models, including B-coefficients, odds ratios and point scores for the three age groups.

#### Infants

The use of LASSO yielded a model comprising of 6 prehospital predictors in infants (Table S4). In a multivariable logistic regression model, male gender, public location, witnessed arrest, having a respiratory cause of arrest, and shockable rhythm were associated with better survival. Having a cardiac etiology and use of prehospital AED were associated with poor survival in this population. This model had an AUROCC of 0.81. We developed point scores by dividing the beta coefficients by the smallest beta (0.26) and rounding the result. The development sample included 9096 infants with 649 (7.1%) surviving to hospital discharge. The validation sample included 3898 infants with 276 (7.1%) surviving to hospital discharge. The performance of the model in development and validation samples are described in Table 2. The model classified 76.5%, 13.5% and 9.8% in low moderate and high survival categories respectively with average probability of survival of 3%, 13% and 34% in the low, moderate and high survival categories. The results were similar for the validation group. Evaluation in the validation group yielded an AUROCC of 0.80.

**Table 2:** Overall comparison of the performance of logistic regression and classification and regression trees.

|  | Logistic regression | Classification and regression trees |
| --- | --- | --- |

|  | Survival/Total (%) |  |  | Survival/Total (%) |  |  |
| --- | --- | --- | --- | --- | --- | --- |
|  | Infants | Preschool<br>and<br>school<br>age | Adolescents | Infants | Preschool<br>and<br>school<br>age | Adolescents |
| **AUROC** |  |  |  |  |  |  |
| Derivation | 0.81 | 0.77 | 0.82 | 0.78 | 0.76 | 0.82 |
| Validation | 0.80 | 0.75 | 0.80 | 0.77 | 0.74 | 0.80 |
| **Number of Predictors** | 6 | 6 | 5 | 3 | 6 | 4 |
| **Survival% by group – Derivation** |  |  |  |  |  |  |
| Low survival | 180/6963<br>(3%) | 36/1385<br>(3%) | 79/1738<br>(5%) | 188/7091<br>(3%) | 93/2410<br>(4%) | 55/1465<br>(4%) |
| Moderate survival | 165/1233<br>(13%) | 408/3297<br>(12%) | 276/1461<br>(19%) | 134/1023<br>(13%) | 141/1254<br>(11%) | 172/1314<br>(13%) |
| High survival | 304/900<br>(34%) | 404/1096<br>(37%) | 434/755<br>(57%) | 327/982<br>(33%) | 614/2114<br>(29%) | 562/1175<br>(48%) |
| **Survival% by group – Validation** |  |  |  |  |  |  |
| Low survival | 85/2980<br>(3%) | 19/574<br>(3%) | 36/755<br>(5%) | 87/3035<br>(3%) | 50/997<br>(5%) | 23/611<br>(4%) |
| Moderate survival | 59/514<br>(11%) | 170/1417<br>(12%) | 83/608<br>(14%) | 50/423<br>(12%) | 56/558<br>(10%) | 62/578<br>(11%) |
| High survival | 132/404<br>(33%) | 179/484<br>(37%) | 166/331<br>(50%) | 139/440<br>(32%) | 262/920<br>(28%) | 200/505<br>(40%) |
| **% of patients in each risk group – Derivation** |  |  |  |  |  |  |
| Low survival | 76.5% | 23.9% | 43.9% | 77.9% | 41.7% | 37.1% |
| Moderate survival | 13.5% | 57.1% | 36.9% | 11.2% | 21.7% | 33.2% |
| High survival | 9.8% | 18.9% | 19.1% | 10.8% | 36.6% | 29.7% |

Abbreviations: AUROC = Area under the receiver operating characteristic curve

#### Preschool and school-age children

In preschool and school-age children, LASSO yielded a model with 6 pre-hospital variables (Table S5). Public location of arrest, witnessed arrest, shockable rhythm, and receiving bystander CPR were associated with better survival. On the other hand, etiology being cardiac or respiratory, and use of AED were associated with poor survival. The model had an AUROCC of 0.77. The development sample included 5778 with 848 (14.7%) surviving to hospital discharge, whereas the validation sample included 2475 children with 368 (14.9%) surviving to hospital discharge. The model classified 23.9%, 57.1% and 18.9% in low, moderate and high survival categories. The average probability of survival was 3%, 12% and 37% for low, moderate and high survival categories, similar for derivation and validation sets. Evaluation in the validation group yielded an AUROCC of 0.75. Model performance characteristics are described in Table 2.

#### Adolescents

In adolescents, LASSO resulted in a model with 5 prehospital predictors (Table S6) and an AUROCC of 0.82. Public location of arrest, witnessed arrest, bystander CPR, and shockable rhythm were associated with improved survival. The model development sample included 3954, out of which 789 (19.9%) survived to hospital discharge, whereas the validation sample included 1694, out of which 285 (16.8%) survived to hospital discharge. The model performance is described in Table 2. This model classified 43.9% in low, 36.9% in moderate and 19.1% in high survival categories with approximately 5%, 19% and 57% average probability of survival in each category respectively. The AUROCC of validation sample was 0.80.

### CART models

#### Infants

The model development sample included 9096 infants with 649 (7.1%) surviving to hospital discharge and yielded an AUROCC of 0.78 using 4 splits and 5 terminal nodes (Figure 1). This model used 3 pre-hospital variables: witnessed arrest, etiology, and first rhythm to classify patients in low, moderate, and high survival categories. Witnessed arrest was considered the most important predictor variable. We manually pruned the tree to remove nodes that did not have clinically meaningful differences in survival between paired terminal nodes. We combined the terminal nodes to create categories for low (<=4%), moderate (5 to 13%), and high (>= 14%) likelihood of survival. The model classified 77.9%, 11.2%, and 10.8% of patients in low, moderate, and high survival categories with an average probability of survival of 3%, 13% and 33% in each category, respectively (Table 2). Evaluating the CART model in the validation sample yielded an AUROCC of 0.77.

**Figure 1:**
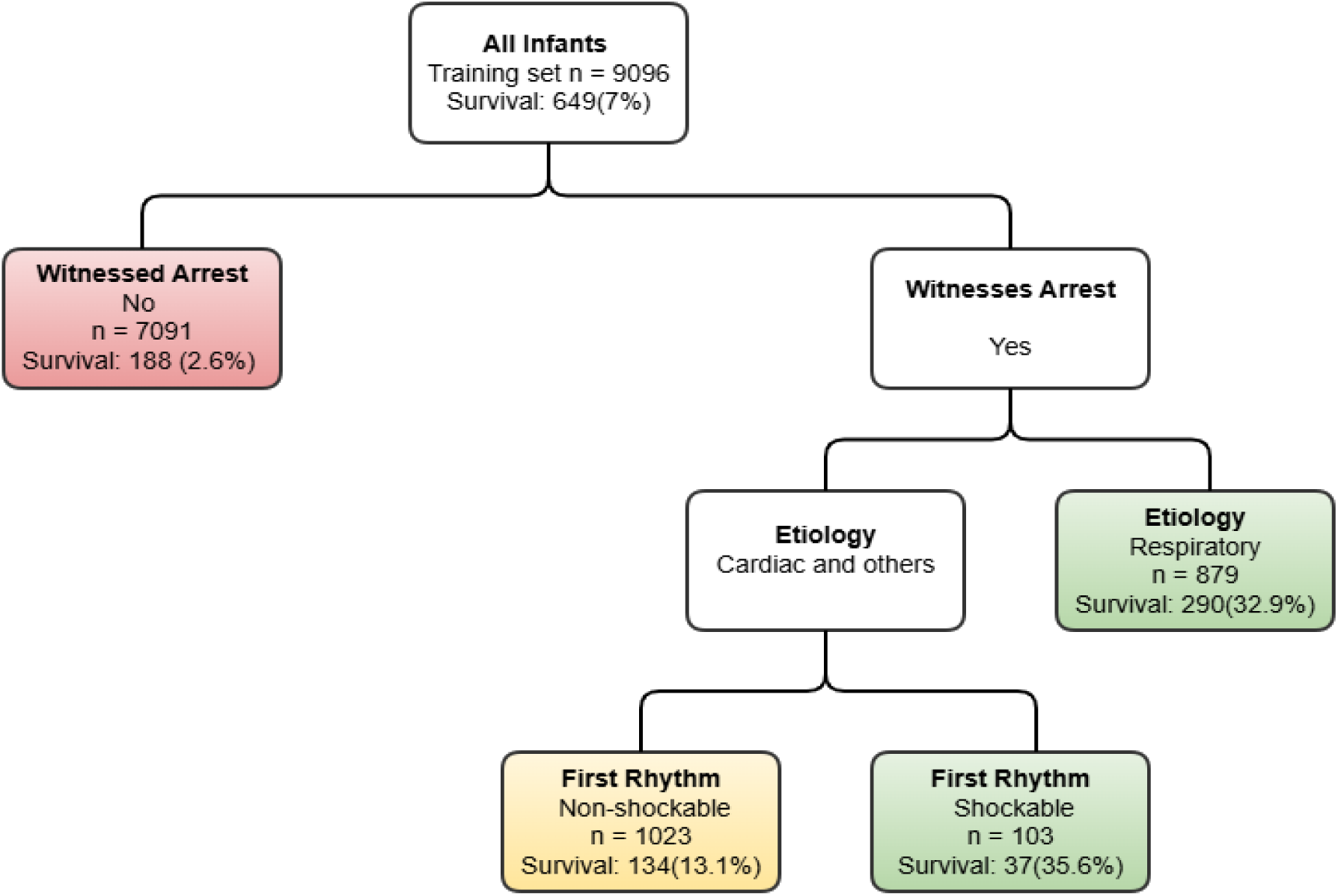
Unsupervised CART models for out-of-hospital cardiac arrest, estimating survival to discharge in infants (development sample n = 9096, validation sample, n = 3898)

#### Preschool and school-age children

The model development sample consisted of 5778 preschool and school-age children with 848 (14.7%) surviving to hospital discharge and yielded an AUROCC of 0.76 using 8 splits and 9 terminal nodes (Figure 2). The model used 6 prehospital variables: witnessed arrest, etiology of arrest, first rhythm, location of arrest, bystander CPR, and prehospital AED use to classify patients in low, moderate, and high survival categories. The most important predictor variable was witnessed arrest. We combined terminal nodes to create categories for low (<= 4%), moderate (5% to 13%), and high likelihood of survival (>=14%). The model classified 41.7%, 21.7%, and 36.6% of patients in the low, moderate, and high survival categories with an average probability of survival of 4%, 11%, and 29% in each category, respectively (Table 2). Evaluating the CART model in the validation sample yielded an AUROCC of 0.74.

**Figure 2:**
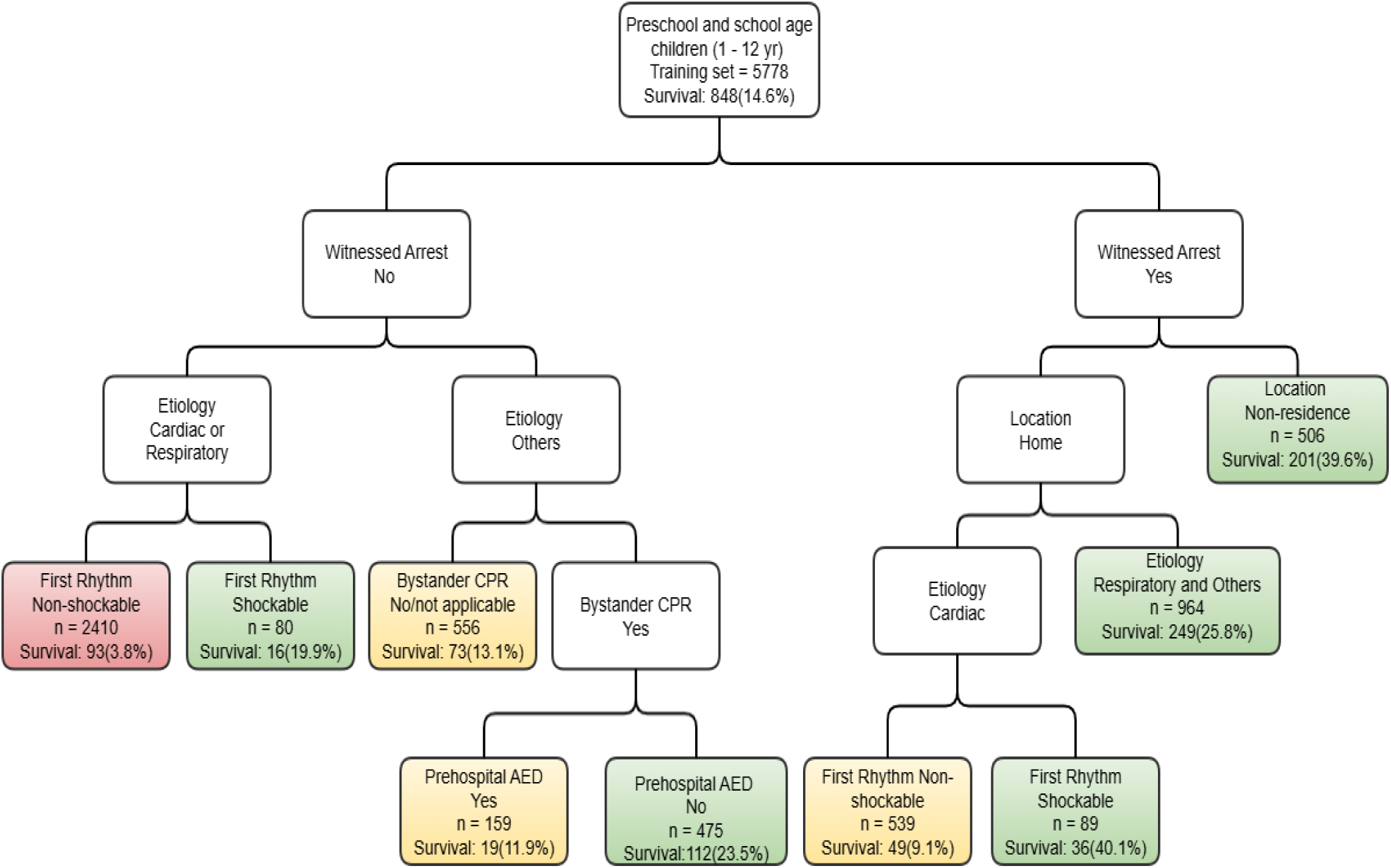
Unsupervised CART models for out-of-hospital cardiac arrest, estimating survival to discharge in preschool and school age children (development sample, n = 5778, validation sample, n = 2475)

**Figure 3:**
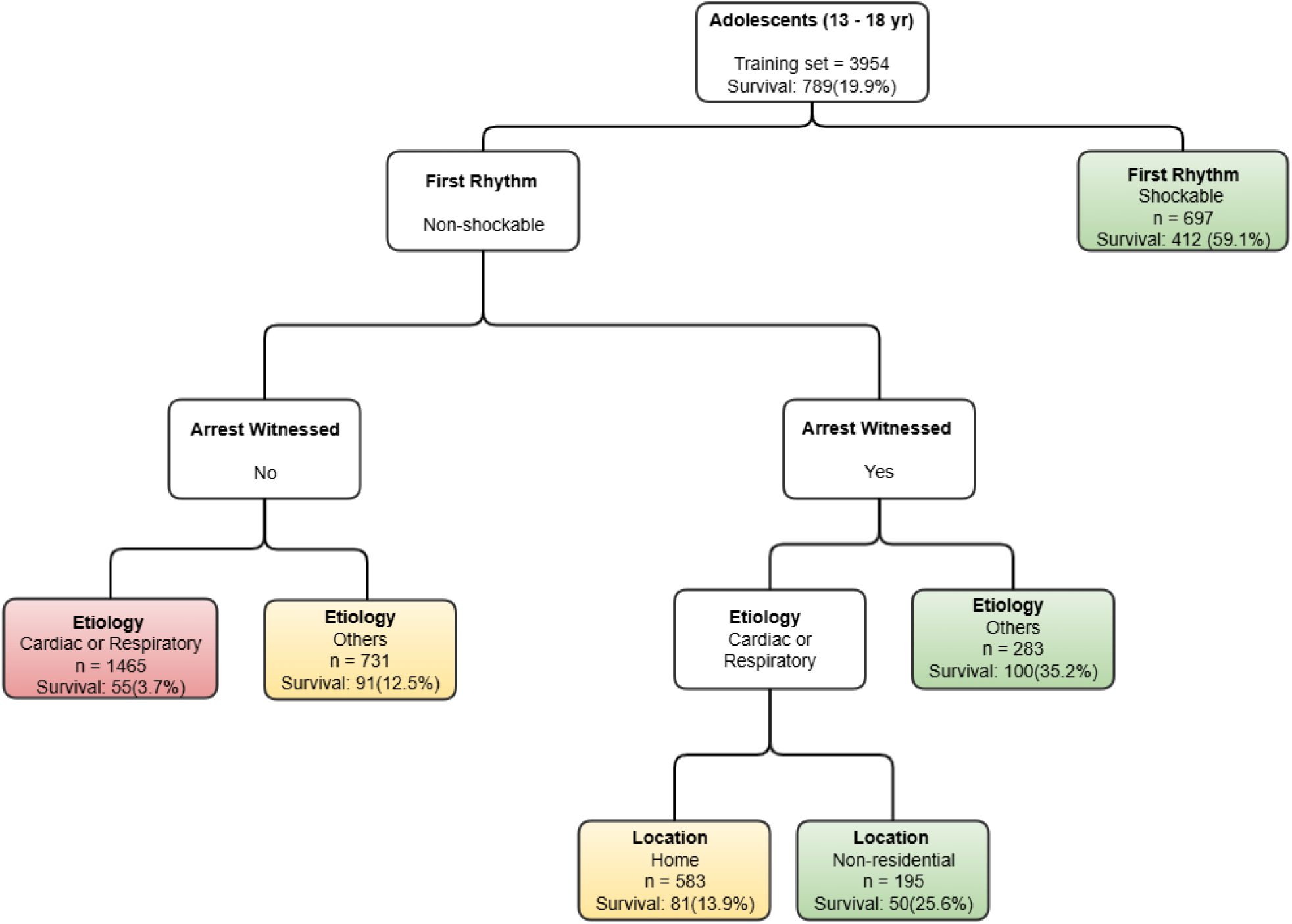
Unsupervised CART models for out-of-hospital cardiac arrest, estimating survival to discharge in adolescents (development sample, n = 3954, validation sample, n = 1694)

#### Adolescents

The model for the development sample consisted of 3954 adolescents with 789 (19.9%) surviving to hospital discharge. The model used 5 splits and 6 terminal nodes and yielded an AUROCC of 0.82. It used 4 prehospital variables: first rhythm, witnessed arrest, etiology of arrest and location of arrest with first rhythm being considered the most important predictor by the algorithm. The terminal nodes were combined to create low (<= 4%), moderate (5% to 16%), and high (>= 17%) survival categories. The model classified 37.1%, 33.2%, and 29.7% of patients in low, moderate, and high survival groups and had an average survival probability of 4%, 13%, and 48% in each group, respectively (Table 2). Evaluating the CART model in the validation sample yielded an AUROCC of 0.80.

## Discussion

We developed and internally validated simple risk scores to predict survival to discharge in pediatric out-of-hospital cardiac arrest patients using logistic regression and CART models. We developed separate models for infants, preschool and school-age children, and adolescents that help classify them into low, moderate, and high survival categories. We found both logistic regression and CART models to be useful. However, several advantages made CART models slightly better than logistic regression models in our study. First, CART models are simpler to implement because of their flowchart representation that does not require calculations. Secondly, the CART models classified more patients in the low and high survival categories compared to logistic regression models that classified more patients in the moderate survival category. Third, CART models used a smaller number of predictor variables compared to logistic regression. Lastly, CART models had accuracy comparable to logistic regression models. Logistic regression models, on the other hand, provide simple point scores that can be easy to implement in research settings. Logistic regression models also have a low variance and the model predictions are relatively stable when applied to different subsets of the same data set or used on an entirely new data set ^16,28–30^.

Both of these sets of risk scores can provide quick and inexpensive estimates of the probability of survival. They can be of great value in helping stratify patients for clinical trials by identifying patients in particular risk categories who may be more or less likely to benefit from a specific intervention. It can help facilitate research across institutions by improving the homogeneity of patient groups and can help develop evidence-based guidelines. It can also help monitor the quality of medical services and may be helpful in clinical decision making by complementing opinion and intuition of a clinician^13–15,31^..

Our results in terms of predictors of survival are largely consistent with previous similar studies from the United States^2,32,33^. Survival was slightly better in our study compared to studies using data from previous years^2^.

This research has several strengths, to the best of our knowledge, this is the first study to develop a risk score to predict the outcomes of pediatric out-of-hospital cardiac arrests using readily available prehospital variables from the nationwide population based registry on OHCA. Our study sample was largely representative of the country’s population. We developed separate models for the three age groups considering inherent differences in survival and predictors associated with survival for these age groups.

This study has a few limitations. We did not have access to pre-arrest labs, sociodemographic and other factors that may be important predictors of survival. We measured survival to discharge and we do not have information about the long-term (3 months, 6 months, 1 year) survival of these participants. Finally, there is a need to prospectively validate these models in a different population.

## Conclusion

We have successfully developed and internally validated risk prediction scores for infants, preschool and school-age children and adolescents with good accuracy. This tool can be helpful in stratifying patients for research and quality improvement projects.

## Data Availability

Data will be made available upon a reasonable request

## Data Sharing Statement

Data will be available on a reasonable request

